## Supplementary File 1 for "HEART rate variability biofeedback for LOng Covid symptoms (HEARTLOC): protocol for a feasibility study"

**Modified COVID-19 Yorkshire Rehabilitation Scale (C19-YRSm)**

**Self-report version**

Patient name:

Hospital number:

Date:	Time:

*The purpose of this questionnaire is to find out more about your current problems following COVID-19 illness. Your responses will be recorded in your clinical notes. We will use this information to monitor your symptoms, offer treatments and assess your response to treatment.*

*This questionnaire will take around 10 minutes. If there are any topics you don’t want to talk about you can choose not to respond.*

*Do you consent for this information to be used for audit and research as well?* **Yes** ☐ **No** ☐

**SYMPTOM SEVERITY**

| *Please answer the questions below to the best of your knowledge.*  ***‘Now’*** *refers to how you feel now/this week (last 7 days).*  ***“Pre-COVID”*** *refers to how you were feeling prior to contracting the illness.*  *If you are unable to recall this, just state ‘don’t know’*  *Rate the severity of each problem on a scale of 0-3:*  ***0 = None; no problem***  ***1 = Mild problem; does not affect daily life***  ***2 = Moderate problem; affects daily life to a certain extent***  ***3 = Severe problem; affects all aspects of daily life; life-disturbing*** | | | |
| --- | --- | --- | --- |
| 1. Breathlessness | Breathlessness: | **Now** | **Pre-COVID** |
|  | a) At rest | **0** ☐ **1** ☐ **2** ☐ **3** ☐ | **0** ☐ **1** ☐ **2** ☐ **3** ☐ |
|  | b) Changing position e.g. from lying to sitting or sitting to lying | **0** ☐ **1** ☐ **2** ☐ **3** ☐ | **0** ☐ **1** ☐ **2** ☐ **3** ☐ |
|  | c) On dressing yourself | **0** ☐ **1** ☐ **2** ☐ **3** ☐ | **0** ☐ **1** ☐ **2** ☐ **3** ☐ |
|  | d) On walking up a flight of stairs | **0** ☐ **1** ☐ **2** ☐ **3** ☐ | **0** ☐ **1** ☐ **2** ☐ **3** ☐ |
| 2. Cough/ throat sensitivity/ voice change | Cough/ throat sensitivity | **0** ☐ **1** ☐ **2** ☐ **3** ☐ | **0** ☐ **1** ☐ **2** ☐ **3** ☐ |
|  | Change of voice | **0** ☐ **1** ☐ **2** ☐ **3** ☐ | **0** ☐ **1** ☐ **2** ☐ **3** ☐ |
| 3. Fatigue (tiredness not improved by rest) | Fatigue levels in your usual activities | **0** ☐ **1** ☐ **2** ☐ **3** ☐ | **0** ☐ **1** ☐ **2** ☐ **3** ☐ |
| 4. Smell/taste | Altered smell | **0** ☐ **1** ☐ **2** ☐ **3** ☐ | **0** ☐ **1** ☐ **2** ☐ **3** ☐ |
|  | Altered taste | **0** ☐ **1** ☐ **2** ☐ **3** ☐ | **0** ☐ **1** ☐ **2** ☐ **3** ☐ |
| 5. Pain/discomfort | Chest pain | **0** ☐ **1** ☐ **2** ☐ **3** ☐ | **0** ☐ **1** ☐ **2** ☐ **3** ☐ |
|  | Joint pain | **0** ☐ **1** ☐ **2** ☐ **3** ☐ | **0** ☐ **1** ☐ **2** ☐ **3** ☐ |
|  | Muscle pain | **0** ☐ **1** ☐ **2** ☐ **3** ☐ | **0** ☐ **1** ☐ **2** ☐ **3** ☐ |
|  | Headache | **0** ☐ **1** ☐ **2** ☐ **3** ☐ | **0** ☐ **1** ☐ **2** ☐ **3** ☐ |
|  | Abdominal pain | **0** ☐ **1** ☐ **2** ☐ **3** ☐ | **0** ☐ **1** ☐ **2** ☐ **3** ☐ |
| 6. Cognition | Problems with concentration | **0** ☐ **1** ☐ **2** ☐ **3** ☐ | **0** ☐ **1** ☐ **2** ☐ **3** ☐ |
|  | Problems with memory | **0** ☐ **1** ☐ **2** ☐ **3** ☐ | **0** ☐ **1** ☐ **2** ☐ **3** ☐ |
|  | Problems with planning | **0** ☐ **1** ☐ **2** ☐ **3** ☐ | **0** ☐ **1** ☐ **2** ☐ **3** ☐ |
| 7. Palpitations/ dizziness | Palpitations in certain positions, activity or at rest | **0** ☐ **1** ☐ **2** ☐ **3** ☐ | **0** ☐ **1** ☐ **2** ☐ **3** ☐ |
|  | Dizziness in certain positions, activity or at rest | **0** ☐ **1** ☐ **2** ☐ **3** ☐ | **0** ☐ **1** ☐ **2** ☐ **3** ☐ |
| 8. Post-exertional malaise (worsening of symptoms) | Crashing or relapse hours or days after physical, cognitive or emotional exertion | **0** ☐ **1** ☐ **2** ☐ **3** ☐ | **0** ☐ **1** ☐ **2** ☐ **3** ☐ |
| 9. Anxiety/ mood | Feeling anxious | **0** ☐ **1** ☐ **2** ☐ **3** ☐ | **0** ☐ **1** ☐ **2** ☐ **3** ☐ |
|  | Feeling depressed | **0** ☐ **1** ☐ **2** ☐ **3** ☐ | **0** ☐ **1** ☐ **2** ☐ **3** ☐ |
|  | Having unwanted memories of your illness or time in hospital | **0** ☐ **1** ☐ **2** ☐ **3** ☐ | **0** ☐ **1** ☐ **2** ☐ **3** ☐ |
|  | Having unpleasant dreams about your illness or time in hospital | **0** ☐ **1** ☐ **2** ☐ **3** ☐ | **0** ☐ **1** ☐ **2** ☐ **3** ☐ |
|  | Trying to avoid thoughts or feelings about your illness or time in hospital | **0** ☐ **1** ☐ **2** ☐ **3** ☐ | **0** ☐ **1** ☐ **2** ☐ **3** ☐ |
| 10. Sleep | Sleep problems, such as difficulty falling asleep, staying asleep or oversleeping | **0** ☐ **1** ☐ **2** ☐ **3** ☐ | **0** ☐ **1** ☐ **2** ☐ **3** ☐ |

**FUNCTIONAL ABILITY**

| 11. Communication | Difficulty with communication/word finding difficulty/understanding others | **Now** | **Pre-COVID** |
| --- | --- | --- | --- |
|  |  | **0** ☐ **1** ☐ **2** ☐ **3** ☐ | **0** ☐ **1** ☐ **2** ☐ **3** ☐ |
| 12. Walking or moving around | Difficulties with walking or moving around | **0** ☐ **1** ☐ **2** ☐ **3** ☐ | **0** ☐ **1** ☐ **2** ☐ **3** ☐ |
| 13. Personal care | Difficulties with personal tasks such as using the toilet or getting washed and dressed | **0** ☐ **1** ☐ **2** ☐ **3** ☐ | **0** ☐ **1** ☐ **2** ☐ **3** ☐ |
| 14. Other activities of Daily Living | Difficulty doing wider activities, such as household work, leisure/sporting activities, paid/unpaid work, study or shopping | **0** ☐ **1** ☐ **2** ☐ **3** ☐ | **0** ☐ **1** ☐ **2** ☐ **3** ☐ |
| 15. Social role | Problems with socialising/interacting with friends* or caring for dependants  *related to your illness and not due to social distancing/lockdown measures | **0** ☐ **1** ☐ **2** ☐ **3** ☐ | **0** ☐ **1** ☐ **2** ☐ **3** ☐ |

**OTHER SYMPTOMS**

Please select any of the following symptoms you have experienced since your illness in the last 7 days. Please also select any previous problems that have worsened for you following your illness.

☐ Fever

☐ Skin rash/ discolouration of skin

☐ New allergy such as medication, food etc

☐ Hair loss

☐ Skin sensation (numbness/tingling/itching/nerve pain)

☐ Dry eyes/ redness of eyes

☐ Swelling of feet/ swelling of hands

☐ Easy bruising/ bleeding

☐ Visual changes

☐ Difficulty swallowing solids

☐ Difficulty swallowing liquids

☐ Balance problems or falls

☐ Weakness or movement problems or coordination problems in limbs

☐ Tinnitus

☐ Nausea

☐ Dry mouth/mouth ulcers

☐ Acid Reflux/heartburn

☐ Change in appetite

☐ Unintentional weight loss

☐ Unintentional weight gain

☐ Bladder frequency,urgency or incontinence

☐ Constipation, diarrhoea or bowel incontinence

☐ Change in menstrual cycles or flow

☐ Waking up at night gasping for air (also called sleep apnea)

☐ Thoughts about harming yourself

Other symptoms – free text

**OVERALL HEALTH**

How good or bad is your health overall in the last 7 days?

For this question, a score of 10 means the BEST health you can imagine. 0 means the WORST health you can imagine.

a) Now:  WORST HEALTH  0 ☐ 1 ☐ 2 ☐ 3 ☐ 4 ☐ 5 ☐ 6 ☐ 7 ☐ 8 ☐ 9 ☐ 10 ☐ BEST HEALTH

b) Pre-Covid:  WORST HEALTH  0 ☐ 1 ☐ 2 ☐ 3 ☐ 4 ☐ 5 ☐ 6 ☐ 7 ☐ 8 ☐ 9 ☐ 10 ☐ BEST HEALTH

**EMPLOYMENT**

Occupation: ______________________

Has your COVID-19 illness affected your work?

☐ No change

☐ On reduced working hours

☐ On sickness leave

☐ Changes made to role/ working arrangements (such as working from home or lighter duties)

☐ Had to retire/ change job

☐ Lost job

Any other comments/concerns:_____________________________________________

**PARTNER/FAMILY/CARER PERSPECTIVE**

This is space for your partner, family or carer to add anything from their perspective:
