## Supplementary File 2a for "HEART rate variability biofeedback for LOng Covid symptoms (HEARTLOC): protocol for a feasibility study"

**The aAP diary sheet**

**Participant Initials	Date**

***Food or fluid intake** – please state what food or drink, including alcohol, was consumed

***Activity** (can be physical, cognitive or emotional) – please state what was the activity and for how long

| **Enter time** | **Position/Activity** | **Blood Pressure** | **Heart**  **Rate** | **Symptoms** | **Other details** |
| --- | --- | --- | --- | --- | --- |
| **EARLY MORNING (ON WAKING)** Time: _ _ hr _ _min | | | | | |
| _ _ hr _ _ min | Lying | _ _ _/ _ _ _  sys. diast |  |  |  |
| _ _ hr _ _min | After 3 min sitting |  |  |  |  |
| _ _ hr _ _min | After 3 min standing |  |  |  |  |
| **BREAKFAST** Time: _ _ hr _ _min; Details of food/fluid*: | | | | | |
| _ _ hr _ _ min | Lying |  |  |  |  |
| _ _ hr _ _ min | After 3 min standing |  |  |  |  |
| **ACTIVITY** Time: _ _ hr _ _min; Details of activity*: | | | | | |
| _ _ hr _ _ min | Before activity |  |  |  |  |
| _ _ hr _ _ min | After 3 min activity |  |  |  |  |
| **LUNCH** Time: _ _ hr _ _min; Details of food/fluid*: | | | | | |
| _ _ hr _ _ min | Lying |  |  |  |  |
| _ _ hr _ _ min | After 3 min standing |  |  |  |  |
| **ACTIVITY** Time: _ _ hr _ _min; Details of activity*: | | | | | |
| _ _ hr _ _ min | Before activity |  |  |  |  |
| _ _ hr _ _ min | After 3 min activity |  |  |  |  |
| **DINNER** Time: _ _ hr _ _min; Details of food/fluid *: | | | | | |
| _ _ hr _ _ min | Lying |  |  |  |  |
| _ _ hr _ _ min | After 3 min standing |  |  |  |  |
| **BEFORE SLEEPING (IN BED)** Time: _ _ hr _ _min | | | | | |
| 22.15pm  (**In bed)** | Lying in usual sleeping position (as with pillows) |  |  |  |  |

**Measure sitting BP/HR only if you find it difficult to stand.**

**Please record any other type of activity that you would like to tell us about and is not listed above, with time & position.**

| **Enter time** | **Position/Activity** | **Blood Pressure** | **Heart**  **Rate** | **Symptoms** | **Other details** |
| --- | --- | --- | --- | --- | --- |
| _ _ hr _ _ min |  |  |  |  |  |
| _ _ hr _ _ min |  |  |  |  |  |
