## Supplementary File 2b for "HEART rate variability biofeedback for LOng Covid symptoms (HEARTLOC): protocol for a feasibility study"

**Adapted Autonomic Profile (aAP) protocol**

**What does it entail?**

Measuring blood pressure (BP) and heart rate (HR) at key times as outlined below while at home, with a personal approved home BP/HR monitor. An example is Omron, approved by the British Hypertension Society. The recordings provide information on how your autonomic nervous system responds to key activities in daily life such as postural change, before/after food and exertion. Experience over the decades indicates that it provides adequate data for initial diagnosis and for guidance on treatment.

Please record time, position, BP and HR, and key symptoms (such as dizziness) in brief on the accompanying **aAP** **diary sheet**. This is of particular importance in autonomic conditions and differs substantially from BP/HR recordings commonly used for high BP. Recordings should be taken on waking, after meals, after exertion and before sleep as outlined below:

- **WAKING** - Take a measurement lying, then after 3 minutes of sitting, then after 3 minutes of standing.
- **EATING** - After a standard meal (breakfast, lunch or dinner), within 10-15 minutes, take a measurement lying, then after 3 minutes of standing. Please note down what food and drink you have consumed (including alcohol) in the space provided.
- **EXERTION** - take a measurement After 3-5 mins of activity (physical, cognitive or emotional) morning and afternoon, separated from lunch and dinner. **NB:** exercise exertion levels will be different for everyone, an example of **physical exertion** can be 5 minutes of gentle walking, or up and down a flight of stairs. An example of **emotional exertion** might be watching an exciting sporting match or film. **Cognitive exertion** might be 5 minutes working out a crossword puzzle. We prefer everyone to attempt at least one form of physical exertion if possible. Please discuss with the clinical team what form of exercise or exertion may be most appropriate for you.

Note that the only reading which we would like you to do seated is the waking reading – a measurement initially on lying, then at sitting for 3 minutes and then at standing for 3 minutes. If it is difficult to stand at other times substitute sitting for standing, especially if after exertion or food.

If you wish to add additional activities, which worsens your symptoms, record them with the time, event/activity and position (lying, sitting, standing).

**Does it involve preparation?**

Ensure that you choose a day when you can complete all of the measurements on the record sheet. It is intended to provide relevant autonomic information during a standard day with usual activities, and thus no change in schedule is needed. The aAP can be repeated on another day if needed.

**What are the advantages of doing the test ?**

The test will inform the clinician about the response of your autonomic system to some of the common triggers or stimuli in daily life. The test also helps you understand what makes your autonomic symptoms worse, which might help you modify some of these activities or triggers.

**Are there any risks of doing the test?**

There is a chance that standing may cause dizziness or even fainting for some people, so please ensure you are leaning against a wall when checking BP/HR on standing. If possible, have another person present in the room whilst performing the test standing. Abandon the test and sit or lie down if symptoms are worse.

**Does it cause discomfort, and are there after-effects?**

The BP cuff may feel uncomfortably tight for a short period if you have a high BP, as some may do, especially while lying down. There should be no after-effects.

**Where does it take place?**

The test can be undertaken in your own home and independent of the GP surgery or hospital. This avoids travel and can be performed whenever convenient. And it can be repeated to determine the effects of treatment.

**How is the result/event sheet forwarded?**

Please enter the results along with your name/ number and the date in the diary sheet and email or post to the clinician/ service:

Address

***The BP/HR autonomic profile and protocol originally was devised and evaluated for autonomic conditions by Professor Mathias, when he directed and developed the UK National Autonomic Referral Units, at St Mary’s Hospital & the National Hospital for Neurology & Neurosurgery @ Queen Square in London. It has been adapted for home use in this protocol and has been of value in the Coronavirus era and its aftermath.***

**June 2022**
